# The Impact of Legislation on Covid-19 Mortality in a Brazilian Federative Unit was Mediated by Social Isolation

**DOI:** 10.1101/2021.06.16.21259057

**Authors:** Fredi A. Diaz-Quijano, Tatiane Bomfim Ribeiro, Aléxia Viana da Rosa, Rossana Reis, Fernando Aith, Deisy F. L. Ventura

## Abstract

**Background:** This study aimed to estimate the effect of restrictive laws on actual social isolation and COVID-19 mortality. Moreover, we evaluated how community adherence, measured with an index of social isolation, would mediate the lockdown effect on COVID-19 mortality.

**Methods:** This ecological study assessed the legislations published until June 30, 2020, in the Brazilian state of Ceará. We performed a systematic review and classification of restrictive norms and estimated their immediate effect on social isolation, measured by an index based on mobile data, and the subsequent impact on COVID-19 mortality (three weeks later). A mediation analysis was performed to estimate the effect of rigid lockdown on mortality that was explained for effective social isolation.

**Results:** The social isolation index showed an increase of 11.9% (95% CI: 2.9% - 21%) during the days in which a rigid isolation norm (lockdown) was implemented. Moreover, this rigid lockdown was associated with a reduction of 26% (95% CI: 21% - 31%) in the three-week-delayed mortality. We also calculated that the rigid lockdown had the indirect effect, i.e., mediated by adherence to social isolation, of reducing COVID-19 mortality by 38.24% (95% CI: 21.64% to 56.07%). Therefore, the preventive effect of this norm was fully explained by the actual population adherence, reflected in the social isolation index. On the other hand, mandatory mask use was associated with 11% reduction in COVID-19 mortality (95% CI: 8% −13%).

**Conclusions:** We estimated the effect of quarantine regulations on social isolation and evidenced that a rigid lockdown law led to a reduction of COVID-19 mortality in one state of Brazil. In addition, the mandatory masks norm was an additional determinant of the reduction of this outcome.

## INTRODUCTION

Several measures have emerged in response to the COVID-19 pandemic, one of them is lockdown which essentially consists of avoiding crowds and restricting the circulation of people in public places. Studies have shown that this type of restriction is associated with reducing the Covid-19 incidence and mortality [1, 2]. In addition, some works based on mathematical modeling have suggested a high impact of social isolation as a control measure [3, 4].

Brazil was one of the countries most affected by the pandemic. By June 16, 2021, there were more than seventeen million confirmed cases and almost five hundred thousand deaths from COVID-19. Sanitary measures focused on the adoption of quarantine with different degrees of restrictions, including the recommendation of physical distancing and suspension of non-essential activities [5]. Moreover, there was a decentralization of the quarantine coordination across and within the Brazilian federative units. Thus, implementation and population adherence to these rules may have varied widely, making it challenging to evaluate the impact of the legislation.

This study aimed to estimate the effect of restrictive measures laws on social isolation and COVID-19 mortality. Moreover, we evaluated how community adherence, measured with an index of social isolation, would mediate lockdown to reduce mortality from COVID-19.

## METHODS

### Design

This ecological study assessed the effect of the legislations published until June 30, 2020, in the Brazilian state of Ceará. This state has implemented strict isolation measures and has been characterized by presenting a regionalization policy, with a diversity of application and scope of rules [6].

### Study setting background

In Brazil, the first wave began right after the first confirmed cases in Europe, in early 2020. Unlike countries like Italy and France, in Brazil lockdown actions have not been instituted from a national level, by the federal government [1, 7, 8]. Instead, the measures were decentralized, so each state led the control strategies locally (5). The most rigorous restrictive actions were practiced by few federative units [6]. In Ceará, a kind of social isolation, considered rigid (with severe restrictions and population duties), was implemented in the most critically affected municipalities.

### Procedures for survey and legislation classification

This study was nested in the Pandemic Rights project [*Direitos na Pandemia*, in Portuguese] that aims to map the legislation issued by the Brazilian Federation and the States about the COVID-19 pandemic. Four researchers identified the norms and extracted the data from the respective official journals filling an electronic form via Google Forms®. The norms were identified by the presence of any of the following descriptors: “Covid-19” or “covid”, “coronavirus” or “corona”, “pandemic”, “health emergency of international importance” and “health emergency of national importance”, “sars-cov-2”, “Federal Law No. 13,979”, or “13,979”. Among those, we considered eligible the rules on public health measures to prevent the spread of covid-19, that established or renewed quarantine, defined essential services, limited the hours or mode of operation of services, or established social distancing policy.

A double-check was performed by an experienced researcher. Each included legislation was characterized by the name, publication date, and validity of the rule. A qualitative analysis classified the restrictions according to the following categories: suspension of events and concerts; functioning of schools; closing of establishments such as bars and restaurants for on-site consumption, commerce in general (including shopping malls), health stores (defined in the norms as “medical and orthopedic, opticians, podiatry, and occupational therapy”); functioning of industries in general, gyms and sports facilities, churches and religious activities; and beach access.

The concept of “rigid isolation”, described in Ceará State Decree No. 33,574 (May 8, 2020), was used for the highest level of restriction. The rigid isolation norm (rigid lockdown) included the “I - special duty of confinement; II - special duty of protection by persons in the risk group; III - special duty of home stay; IV - control of the circulation of private vehicles; V-control of entry and exit from the municipality”. Additionally, the rule for mandatory facial mask use was also considered as an independent variable.

For the regionalized measures (which did not include the entire state), the coverage was estimated as the proportion of the population affected by the rule in relation to the total of the Ceará state, according to IBGE data with 2020 estimated population [9].

### Epidemiological indicators

We chose mortality as the outcome variable because we considered it the most reliable measure. The above because other indicators, as the number of total cases (including non-fatal), could be more affected by underreporting and might not reflect disease burden trends. We consider this based on what happens with other infectious diseases, in which the surveillance system shows greater sensitivity for identifying the most severe cases, such as deaths [10, 11]. The daily and cumulative counts of deaths by COVID-19 were obtained from the Brazilian Ministry of Health through the electronic domain [12].

To evaluate social isolation, we used the daily social isolation index provided by the InLoco initiative, which is based on mobile data [13]. The technology of Inloco detects when the mobile device remains for long periods in a certain location and sends the location information and the smartphone’s advertising identification number (Advertising ID) to its servers without directly identifying the users. The Isolation Index is calculated as the ratio of users who did not leave their residence place, on a given day, to the total number of users in that same region. Thus, the higher the index, the greater the estimated degree of isolation.

### Data analysis

We considered the analysis units the days of the period between March 16, 2020, and May 07, 2020, and extended the follow-up for epidemiological outcomes until July 26, 2020. Independent variables were created to classify each day according to implementation of each legislation evaluated (exposure variables).

We elaborated a directed acyclic graph (DAG) to guide adjustments during the analyses (Figure 1). We considered, for example, that the implementation of legislation, adherence to social isolation, and COVID-19 mortality, varying with time. Furthermore, baseline disease burden could both affect isolation adherence and be the main determinant of transmission and mortality. In addition, several (unmeasured) determinants could affect both baseline disease burden and isolation adherence (Figure 1).

**Figure 1:**
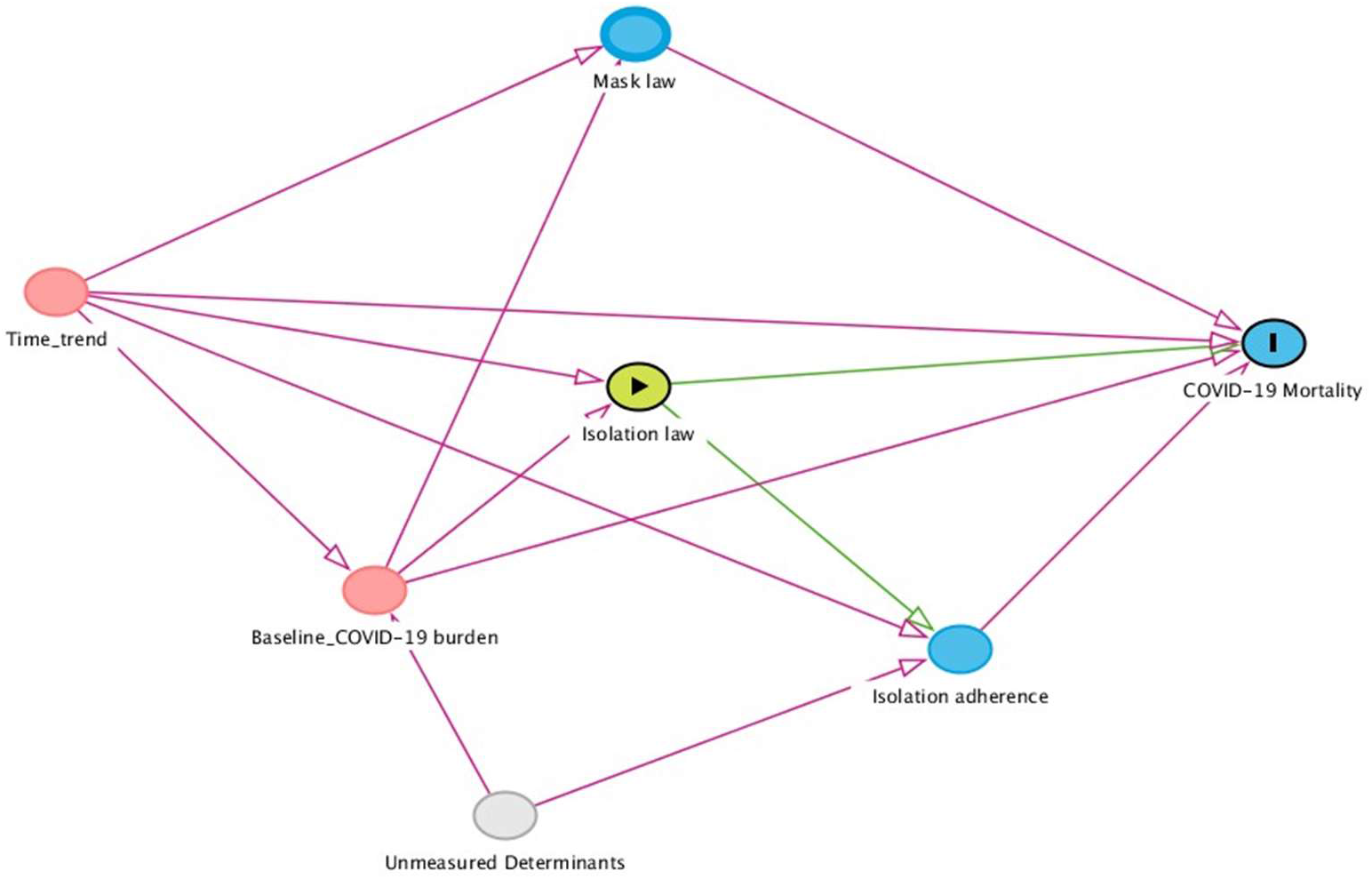
Directed acyclic graph (DAG) of determinants of COVID-19 mortality.

We evaluated the association between each restriction category contemplated by the legislation and the social isolation index recorded on the same days of legislation implementation. This index presented a distribution compatible with a normal one (Figure 2). Thus, we used linear regression to estimate the effects on this variable. Based on the DAG, for these estimations, we adjusted for time and cumulative COVID-19 mortality until the day before the index measurement. Social isolation index was analyzed without delay because it was expected an immediate effect of the legislation.

**Figure 2:**
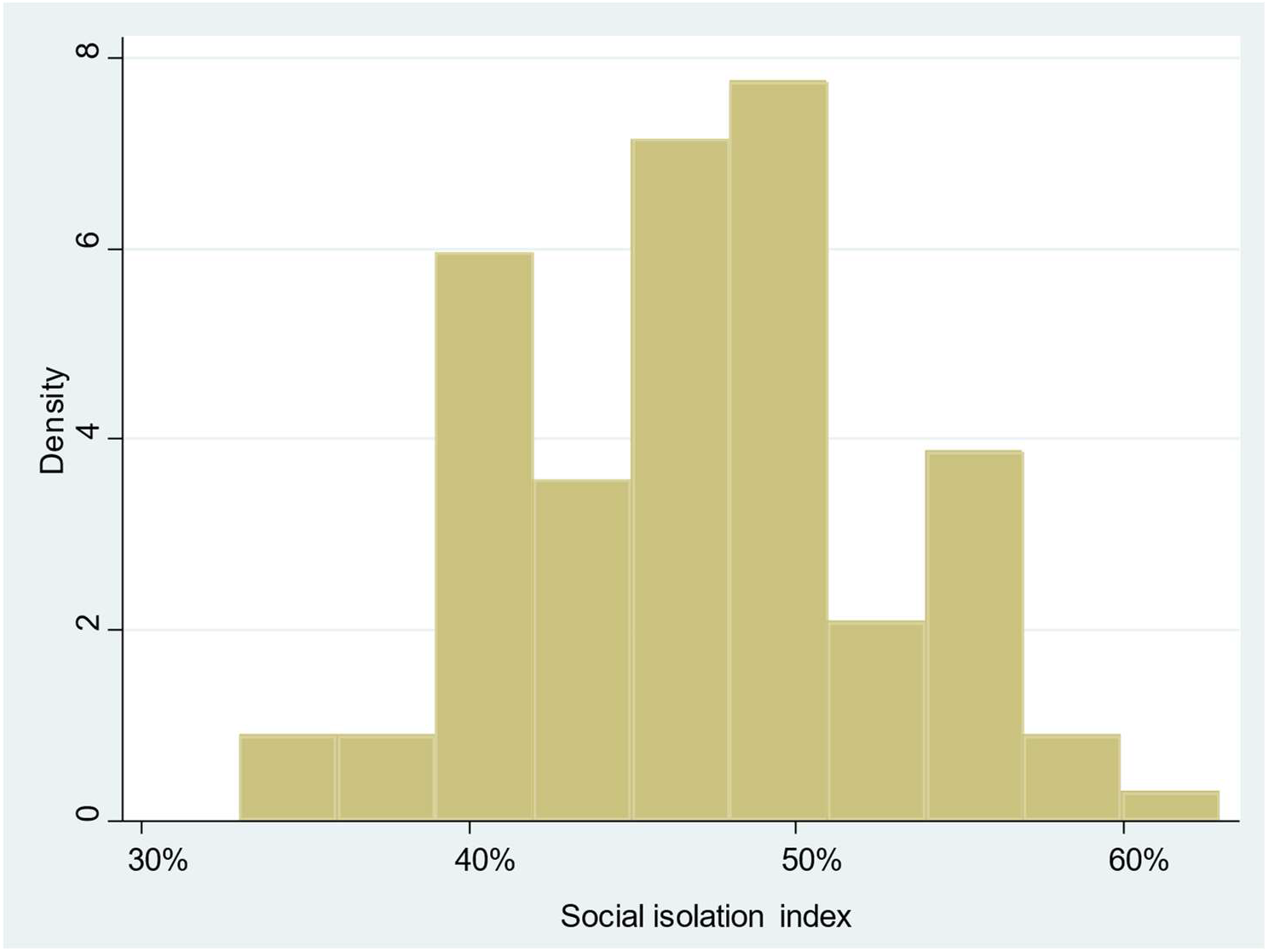
Distribution of the social isolation index in the state of Ceará from January to June 2020.

After, we assessed the legislation effect on the daily death count recorded three weeks later. This interval was chosen considering the likely incubation period and time between symptom onset and death [14, 15]. In addition, we also supported this interval on an exploratory analysis in which we evidenced a relationship between a social event that generated people agglomeration and the subsequent increase in mortality. For this exploratory analysis, we observed new death distribution per week from December 2020 to March 2021 and identified a progressive increment of COVID-19 deaths after the Carnival date, which was especially accentuated three weeks after this traditional holiday (Figure 3).

**Figure 3.**
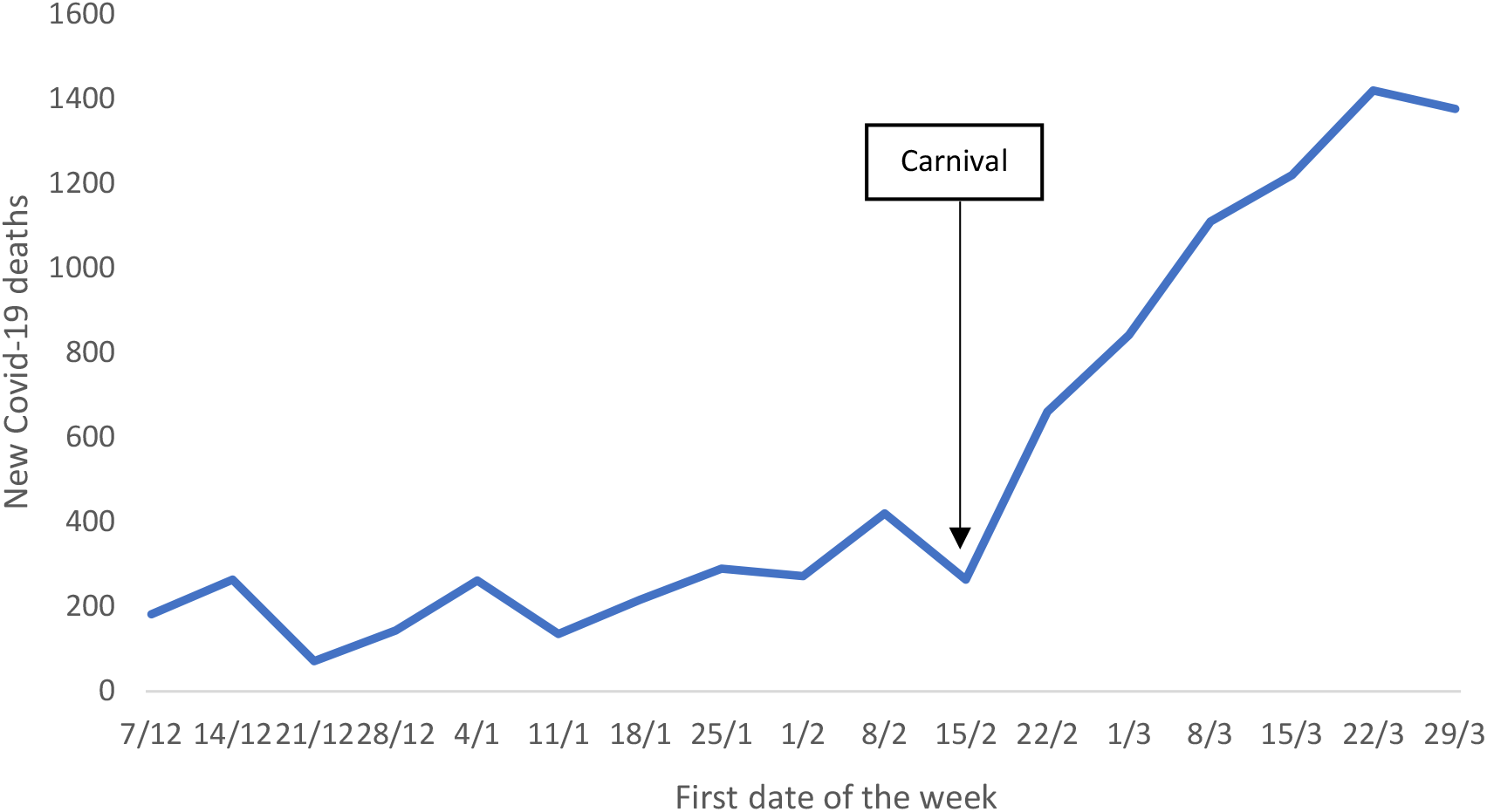
Weekly COVID-19 mortality in Ceará, Brazil, December 2020 to March 2021.

Thus, we used Poisson regression to estimate the effect of the legislation on the subsequent new deaths counted daily (considering the delay of 21 days). This estimation was adjusted for time and former COVID-19 mortality, cumulated up to seven days after the exposure time (baseline disease burden indicator).

Additionally, we assessed how much the effect of the rigid isolation norm (rigid lockdown) on COVID-19 mortality was mediated by population adherence, measured with the social isolation index. For this purpose, the indirect effect was calculated as the product between α, the regression coefficient of the isolation index on the legislation (adjusted for time and cumulative COVID-19 mortality up to the previous day of norm exposure); and β, the (Poisson) regression coefficient of the death count 21 days after the norm exposure on the isolation index (adjusted for the corresponding legislation, time, and cumulative COVID-19 mortality up to seven days after the norm exposure). Namely,

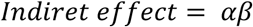

The standard error of this indirect effect, *E*_i_, was calculated as:

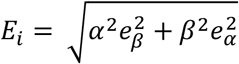

Where *e*_*α*_ e *e*_*β*_ corresponded to the standard errors of α and β, respectively. Based on this, Sobel’s test was performed by calculating its statistic, with Z distribution, as *αβ*/*E_i_*.

We used incidence rate ratios (IRR) as association measures, so both the total and indirect effects were presented as reductions of IRR (1-IRR). STATA v.15.1 software was used for the statistical analyses.

## RESULTS

From January to June 2020, the state of Ceará issued fourteen norms dealing with quarantine measures related to pandemic control, all from the Office of Governor, the first rule dated March 16th, 2020. On May 5th, 2020, Decree No. 33,574, which instituted rigid isolation in the municipality of Fortaleza, was implemented, and it was established the regionalization of policies according to the epidemiological profile of COVID-19 incidence/mortality.

The social isolation index showed an average increase of 11.9% during days of rigid isolation (95% CI: 2.9% - 21%). In addition, other specific restrictions were also associated with a significant increase in the social isolation index, including closing of restaurants/bars, specific healthcare stores, industries, gyms, and restriction of religious activities (Table 1). As expected, mask use norm did not affect this isolation index based on mobility.

**Table 1:** Relationship between restrictive legislation and social isolation index.

| Restrictive measures | Regression coefficient (95%CI)* | p-value |
| --- | --- | --- |
| Rigid isolation norm (lockdown) | 11.93 (2.91 a 20.96) | 0.010 |
| Specific restrictions applied to: |  |  |
| Restaurants/bars | 13.21 (8.62 a 17.81) | <0.001 |
| Schools | -10.52 (-24.29 a 2.79) | 0.12 |
| Specific healthcare stores | 11.19 (8.14 a 14.24) | <0.001 |
| General Commerce | -0.86 (-13.52 a 11.78) | 0.89 |
| Gyms | 15.66 (11.17 a 20.15) | <0.001 |
| Industry | 11.19 (8.13 a 14.24) | <0.001 |
| Beach | 15.66 (11.17 a 20.15) | <0.001 |
| Religious activities | 13.21 (8.62 a 17.81) | <0.001 |
| Mandatory use of facial mask | 2.06 (-14.51 a 5.56) | 0.247 |

\* Increases in percentage units of the social isolation index. All estimates are adjusted for time and cumulative COVID-19 mortality up to the previous day of index measurement.

Rigid lockdown was associated with a reduction of 26% (95% CI: 21% - 31%) in the three-week-delay mortality. The other restrictive measures were not significantly associated with this outcome (Table 2). However, mandatory mask use was associated with 11% reduction in mortality (95% CI: 8% - 13%). When included in the same model, both the rule of rigid isolation and mask use norm, were independently associated with reduction in mortality (IRRs: 0.79, 95%CI: 0.73 - 0.86; and, 0.96, 95%CI: 0.93 - 1, respectively).

**Table 2:** Association between restrictive legislation and COVID-19 mortality recorded three weeks after the norm implementation.

| Restrictive measures | Rate ratio (95%CI) * | p-value |
| --- | --- | --- |
| Rigid isolation norm (lockdown) | 0,74 (0,69-0,79) | <0.001 |
| Specific restrictions applied to: |  |  |
| Restaurants/bars | 0,96 (0,90-1,03) | 0,304 |
| Schools | 0,92 (0,84-1,01) | 0,507 |
| Specific healthcare stores | 0,99 (0,96-1,02) | 0,529 |
| General Commerce | 0,95 (0,88-1,03) | 0,250 |
| Gyms | 1,03 (0,95-1,14) | 0,515 |
| Industry | 0,99 (0,96-1,02) | 0,529 |
| Beach | 1,03 (0,93-1,14) | 0,515 |
| Religious activities | 0,96 (0,90-1,03) | 0,304 |
| Mandatory use of facial mask | 0,89 (0,87-0,92) | <0.001 |

\*Each estimate was adjusted by cumulative deaths up to 7 days after the application of the norm and time (date of the norm).

### Mediation analysis

We estimated that for each 1% increase in the isolation index there was a 4% relative reduction in the mortality rate (95% CI: 3.8% - 4.2%; p<0.001) recorded three weeks later, in a model that adjusted for the rigid isolation legislation, time, and the previous COVID-19 mortality (up to 7 days after the norm application). Moreover, we calculated that the rigid lockdown had the indirect effect, i.e., mediated by adherence to social isolation, of reducing COVID-19 mortality by 38.24% (95% CI: 21.64% to 56.07%, p=0.009). That was, a higher value than the total estimated effect of the legislation. Furthermore, in the model with the mediator, no direct protective effect of the rigid isolation norm was observed. Therefore, the prevention of COVID-19 mortality attributable to this norm was fully explained by the actual population adherence, reflected in the social isolation index.

## DISCUSSION

Rigorously implemented public health policies are reflected in epidemic control [16]. New Zealand is an example of reduction of COVID-19 disease burden, mainly due to effective case isolation and population testing [17]. We estimated the effect that restrictive measures implemented by a local government appear to have on social isolation. As expected, the strongest association was shown with the rigid lockdown which was the only restrictive norm that was significantly associated with a reduction in mortality. Interestingly, this effect would be explained entirely by population adherence (as measured by the social isolation index). Thus, the mediation analysis helped with the documentation of the legislation effect by explaining the association through the expected mechanism.

Other studies have identified a relationship between lockdown implementation and a reduction in mortality [1, 7, 8]. Our analysis was consistent with those reports and additionally presented a quantification of the effect of several restrictive norms on social isolation. Moreover, it contributes a mediation analysis that supports more robust conclusions while highlighting the usefulness of the social isolation index as a real-time indicator of community adherence to issued norms.

When community transmission has been established, social isolation measures correlate with reproductive number and the COVID-19 incidence [18]. In the present study, the social isolation index was directly related to the of regulations, mainly when included intense restrictions on people circulation, closure of gyms and sports venues, bars and restaurants, religious activities, beaches, and industries. Thus, with few exceptions (schools and general commerce), restrictive measures had a fairly consistent effect on social isolation.

The gradual opening of establishments was a conservative conduct in the state of Ceará, in the norms evaluated until June 2020. Only industries were operating in the entire state. Furthermore, schools and religious activities had been relaxed only for the Municipality of Fortaleza, which represents almost 30% of the state population. This gradual release and varied scope of the norms allowed to have several gradients, and we consider that it favored the evaluation of the legislations.

Besides, in this work we observed that the mandatory use of masks makes an additional contribution in reducing mortality. Moreover, the fact that this norm is not associated with the social isolation index is an interesting result because it highlights the specificity of this index as a mediator of quarantine measures. Face masks protect against infection by several respiratory diseases [19]. Modeling studies with the population of two North American cities showed that the use of masks by 80% of the population could reduce mortality by 24-65% [20]. The discrepancy between those estimates and ours could suggest that differing from the modeling results, in practice, the adherence might be lower than desired. However, the documented effect was significant and complementary to the isolation rule. Therefore, the promotion of continued mask use should also be encouraged.

Because this is an ecological study, we do not know the distribution of individual exposures and attitudes. Thus, variations in communication and implementation of norms during the different phases of the pandemic may affect their evaluation. During the observation period, social isolation index ranged from approximately 33% to 63%. This exemplifies the wide range of potential community responses and also the need to reinforce the legislation with motivational messages and complementary policies to support lockdown.

On the other hand, there was no a systematic measure to assess adherence to mask use, so the estimated effect of the norm may be underestimating the potential usefulness of this practice in preventing transmission and, therefore, COVID-19 mortality. In spite of these limitations, this work makes a contribution to support preventive policies with epidemiological evidence. Furthermore, we highlight the usefulness of the social isolation index to monitor adherence to quarantine measures.

## Conclusion

In this study, we estimated the effect of quarantine regulations on the social isolation and evidenced that a rigid lockdown law led to a reduction of COVID-19 mortality in one state of Brazil. In addition, we documented the role of mandatory masks norm as an additional determinant of the reduction of this outcome.

## Data Availability

All the data analyzed are public.

## Funding

CONECTAS Human Rigths [Direitos Humanos], agreement 01/2020. National Council of State Secretaries of Health [Conselho Nacional de Secretários Estaduais de Saúde – Conass], process 080/2020, contract 044/2020. FADQ was granted a fellowship for research productivity from the Brazilian National Council for Scientific and Technological Development – CNPq, process/contract identification: 312656/2019-0.

## Competing interest

No author has conflicts of interest related to this study.

## Notes

### Competing Interest Statement

The authors have declared no competing interest.

### Author Declarations

According to the Brazilian National Health Council [Conselho Nacional de Saude], research using secondary official and public data does not need an Ethics Committee evaluation (Resolution CNS 510/2016, Art. 1, II e III).

